# Structured retrieval closes the gap between low-cost and frontier clinical language models

**DOI:** 10.64898/2026.03.22.26349018

**Authors:** Alon Gorenshtein, Moran Sorka, Mahmud Omar, Keren Miron, Adi Ahituv, Yiftach Barash, Eyal Klang, Shahar Shelly

**Affiliations:** Department of Neurology, Harvard Medical School, Boston, MA, USA; Department of Neurology, Beth Israel Deaconess Medical Center, Boston, MA, USA; BRIDGE GenAI Lab, Beth Israel Deaconess Medical Center, Harvard Medical School, Boston, MA, USA; AI Neurology Laboratory, Ruth and Bruce Rapaport Faculty of Medicine, Technion Institute of Technology, 3525408 Haifa, Israel; Department of Radiology, Beth Israel Deaconess Medical Center, Harvard Medical School, Boston, MA, USA; Department of Neurology, Rambam Health Care Campus, Haifa, Israel; Department of Neurology, Mayo Clinic, Rochester, MN, USA

## Abstract

Most clinical large language model (LLM) benchmarks rely on clean, concise vignettes that do not reflect the noisy, long-form documentation typical of real clinical records. How LLM performance degrades under realistic chart conditions remains poorly characterised. Here we test whether structured retrieval workflows protect National Institutes of Health Stroke Scale (NIHSS) scoring accuracy under systematic context stress. Using 100 de-identified acute stroke cases and a fully crossed 4 x 4 x 3 x 3 condition matrix (144 conditions per case), we vary context acquisition method, document length, distractor load and critical-information position across four Gemini models (57,047 retained runs). Structured retrieval reduces mean absolute error (MAE) from 4.58 to 2.96 points relative to non-agentic baselines (mean gain 1.62 MAE points; 95% CI 1.57–1.67; 35% relative reduction), with consistent gains across all 36 stress combinations. Lower-cost models show disproportionately larger gains (2.76 versus 0.45 MAE points). Tool-retrieved pipelines outperform retrieval-augmented generation in 33 of 36 combinations. These findings indicate that retrieval architecture, rather than model scale alone, is a tractable lever for robust, equitable clinical LLM deployment.

## Introduction

Large language model (LLM) performance is highly contingent on the quality and structure of the input context^1,2^. In clinical practice, models must process messy charts rather than clean vignettes, facing context stress that includes long and heterogeneous notes, variable signal-to-noise ratios and clinically decisive findings that may appear late or inconsistently across the record^3,4^. As clinical documentation expands, records become longer and noisier^5^. Models must infer what to prioritise under competing signals, and small interface or retrieval choices can cause decisive findings to be diluted, displaced or effectively ignored^6^. At scale, this becomes a safety concern because the same failure pattern can repeat across many encounters.

These vulnerabilities are particularly consequential in acute stroke workflows, where errors in National Institutes of Health Stroke Scale (NIHSS) scoring can affect triage, treatment urgency and downstream care pathways^7–9^. The NIHSS is a standardized, 11-item neurological assessment tool used to quantify stroke severity. It evaluates level of consciousness, gaze, visual fields, facial palsy, motor function in the arms and legs, limb ataxia, sensory function, language (aphasia), dysarthria, and extinction/inattention. Each item is scored on an ordinal scale (0–4 for most items), with total scores ranging from 0 to 42; higher scores reflect greater neurological impairment. It has strong interrater reliability when administered by trained examiners and correlates with infarct volume and functional outcome.

Most evaluation paradigms treat clinical context as static input, yet deployed systems increasingly construct context through interface choices, retrieval pipelines and iterative prompting^10^. Controlled studies show that LLM performance depends not only on what evidence is present but on where it appears, with serial-position effects in long inputs and marked degradation when decisive information is buried mid-context^6^. Long-context benchmarks further indicate that nominal context-length claims can overstate effective use, as accuracy drops with increasing length and task complexity^11^. These failures may be amplified when distractors resemble what retrieval systems actually return^12^. At the same time, the field is shifting from single-shot prompting toward agentic workflows that interleave reasoning with tool use^13^, raising a practical design question: do structured retrieval frameworks worsen context stress by adding intermediate text, or do they improve reliability by enabling more targeted information acquisition?

Here we address this question in a controlled NIHSS scoring setting by systematically varying context acquisition method, document length, noise and information position.

## Results

### Overall retrieval-based protection effect

Across 14,256 paired analysis units, structured retrieval workflows reduced MAE from 4.575 to 2.959, corresponding to a retrieval-based protection (RBP) gain of 1.616 MAE points (35.3% relative reduction; 95% CI 1.565–1.669; Fig. 1a).

**Figure 1.**
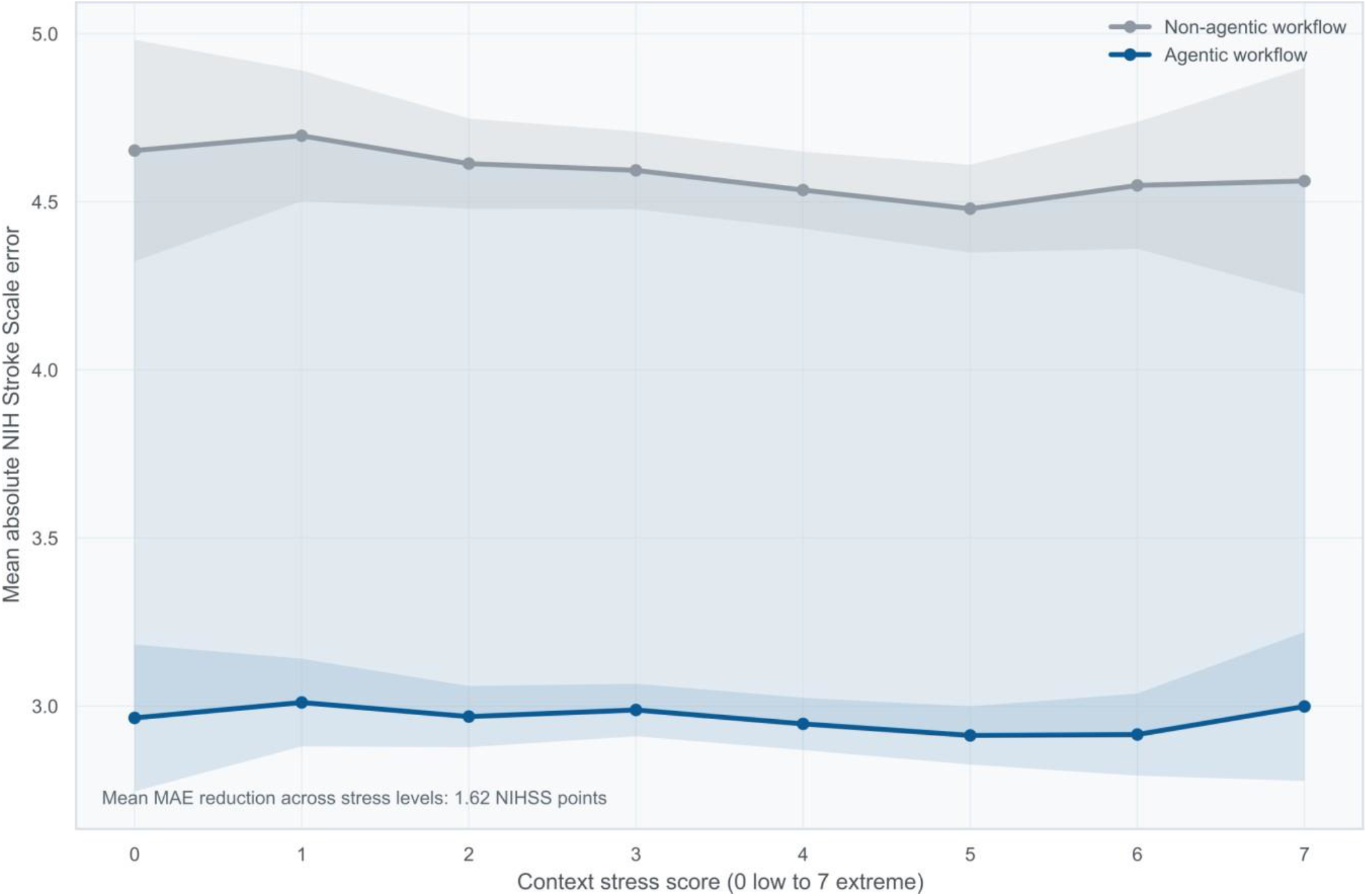
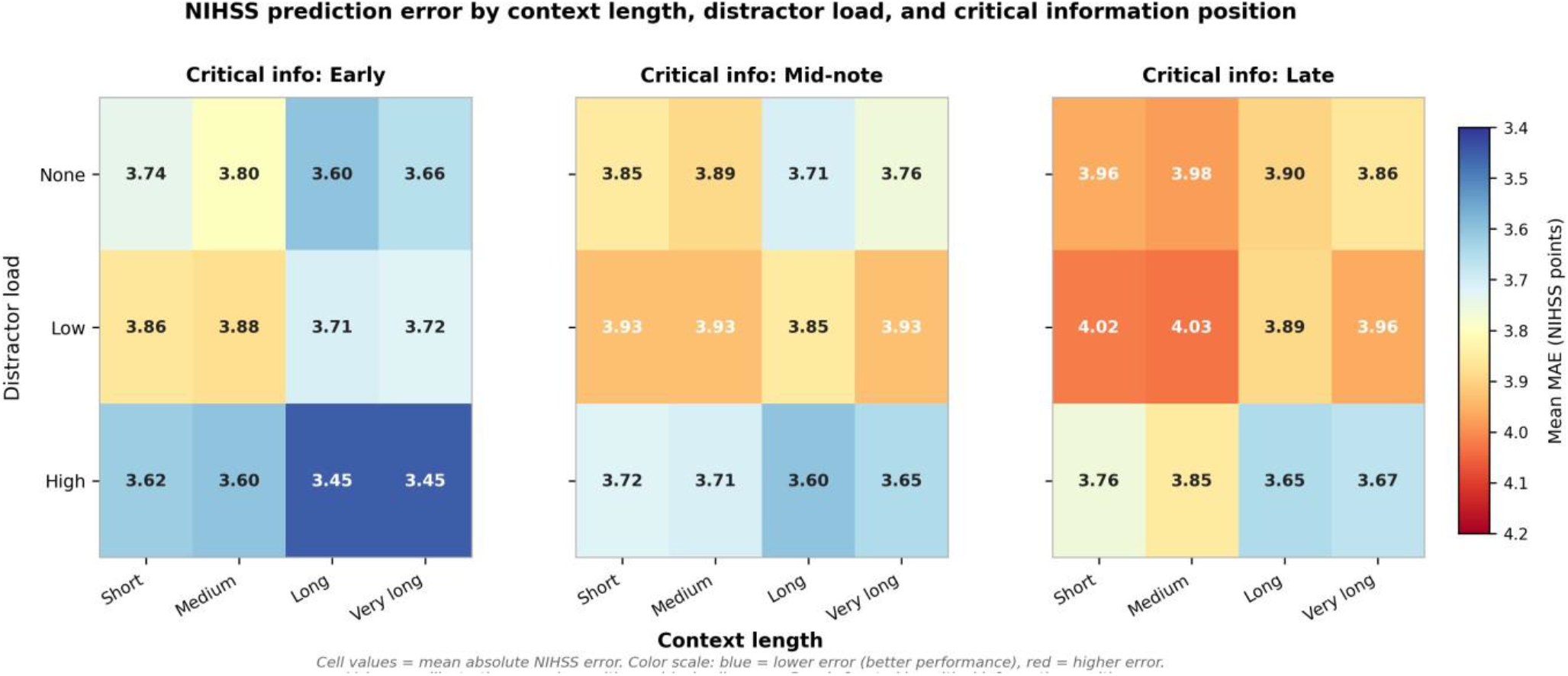
Overall retrieval-based protection effect on NIHSS scoring accuracy. (a) Mean absolute error (MAE) for non-agentic and agentic workflows across composite stress scores (0–7). Shaded bands denote bootstrap 95% CIs (4,000 resamples). The mean RBP gain of 1.62 MAE points was consistent across all stress levels. (b) Faceted heatmap displaying MAE of NIH Stroke Scale score prediction across all combinations of three experimental factors: context length (columns), distractor load (rows), and critical information position (panels). Each cell represents the mean MAE for one factor-level combination. Panels are faceted by critical information position (Early, Mid-note, Late). Distractor load levels reflect the degree of clinically irrelevant information embedded in the input (None, Low, High). Context length categories reflect the total token length of the clinical note (Short, Medium, Long, Very long).

### Protection was concentrated in lower-cost models

In weaker models, MAE decreased from 6.560 to 3.798 (gain 2.762; 95% CI 2.678–2.849; 42.1% relative reduction). In stronger models, MAE decreased from 2.550 to 2.103 (gain 0.447; 95% CI 0.409–0.483; 17.5% relative reduction). The weak-minus-strong gain differential was 2.316 MAE points (95% CI 2.223–2.412).

At the individual-model level, gains were 4.102 (Gemini 2.5 Flash-Lite), 1.423 (Gemini 2.5 Flash), 0.487 (Gemini 3 Pro Preview) and 0.405 (Gemini 3 Flash Preview) (Table 1, Fig. 2).

**Table 1.** Retrieval-based protection gains by model and model class. RBP gain is the paired MAE difference (non-agentic minus agentic). Bootstrap 95% CIs based on 4,000 resamples (seed 42). N, number of paired analysis units.

| Group | N | Non-agentic MAE | Agentic MAE | RBP gain | Reduction (%) | 95% CI low | 95% CI high |
| --- | --- | --- | --- | --- | --- | --- | --- |
| Overall | 14,256 | 4.575 | 2.959 | 1.616 | 35.33 | 1.565 | 1.669 |
| **Weaker models** | 7,200 | 6.560 | 3.798 | 2.762 | 42.11 | 2.678 | 2.849 |
| **Stronger models** | 7,056 | 2.550 | 2.103 | 0.447 | 17.52 | 0.409 | 0.483 |
| Gemini 2.5 Flash-Lite | 3,600 | 9.048 | 4.946 | 4.102 | 45.33 | 3.961 | 4.246 |
| Gemini 2.5 Flash | 3,600 | 4.072 | 2.649 | 1.423 | 34.95 | 1.351 | 1.497 |
| Gemini 3 Pro Preview | 3,600 | 2.827 | 2.341 | 0.487 | 17.21 | 0.426 | 0.548 |
| Gemini 3 Flash Preview | 3,456 | 2.261 | 1.856 | 0.405 | 17.93 | 0.360 | 0.451 |

**Figure 2.**
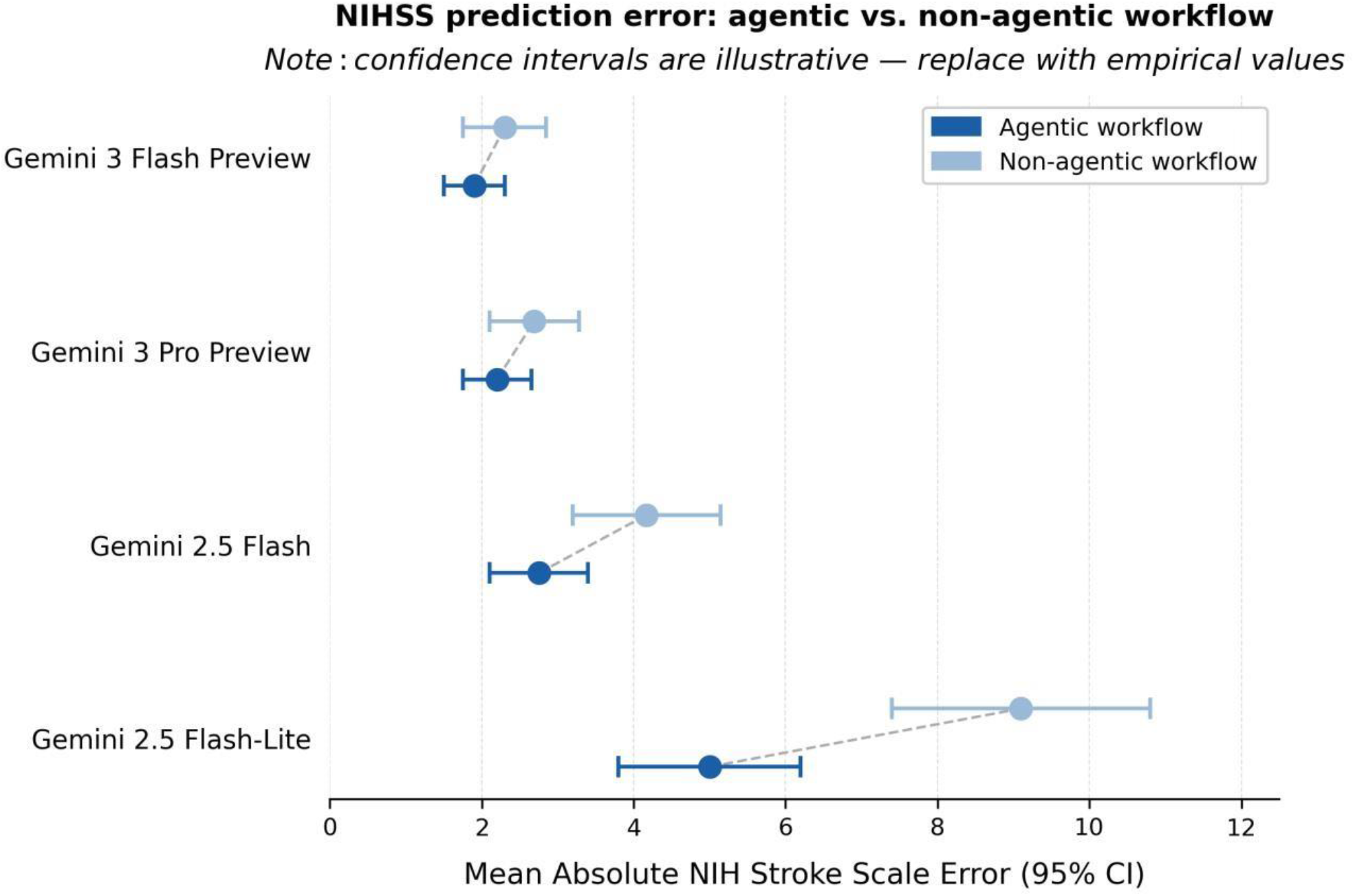
Agentic workflow reduces NIHSS prediction error across all evaluated models. Dot plot showing mean absolute error (MAE) of NIH Stroke Scale score prediction for agentic (dark blue) and non-agentic (light blue) workflows across four large language models. Point estimates with 95% confidence intervals are shown for each model–workflow pair. Models are ordered by non-agentic baseline error. Lower-cost models (Gemini 2.5 Flash-Lite, Gemini 2.5 Flash) demonstrated greater absolute MAE reduction with agentic retrieval than frontier models (Gemini 3 Pro Preview, Gemini 3 Flash Preview), suggesting a larger performance ceiling effect in less capable models. Dashed lines connect paired estimates within each model. Confidence intervals are derived from case-level prediction variance across the evaluation cohort.

### Gains were consistent across stress strata

RBP gains remained positive across all tested strata, including context length (1.575–1.651 MAE), distractor load (1.571–1.702 MAE), information position (1.597–1.653 MAE) and composite stress score (range 0–7; gains 1.563–1.694 MAE). Weak-minus-strong differentials remained large across all stress domains (2.165–2.486 MAE points).

### Condition-level mapping identified residual-risk archetypes

All 36 condition combinations showed positive RBP gain in pooled data and in both model classes. Pooled gains ranged from 1.382 to 1.878 MAE; weak-model gains from 2.295 to 3.353 MAE; and strong-model gains from 0.167 to 0.726 MAE.

The highest-gain combination was long context | high distractor | late position (pooled gain 1.878 MAE; weak-model gain 3.353 MAE). The highest residual-risk combination under structured retrieval was very long context | low distractor | late position (agentic MAE 3.215; Fig. 3).

**Figure 3.**
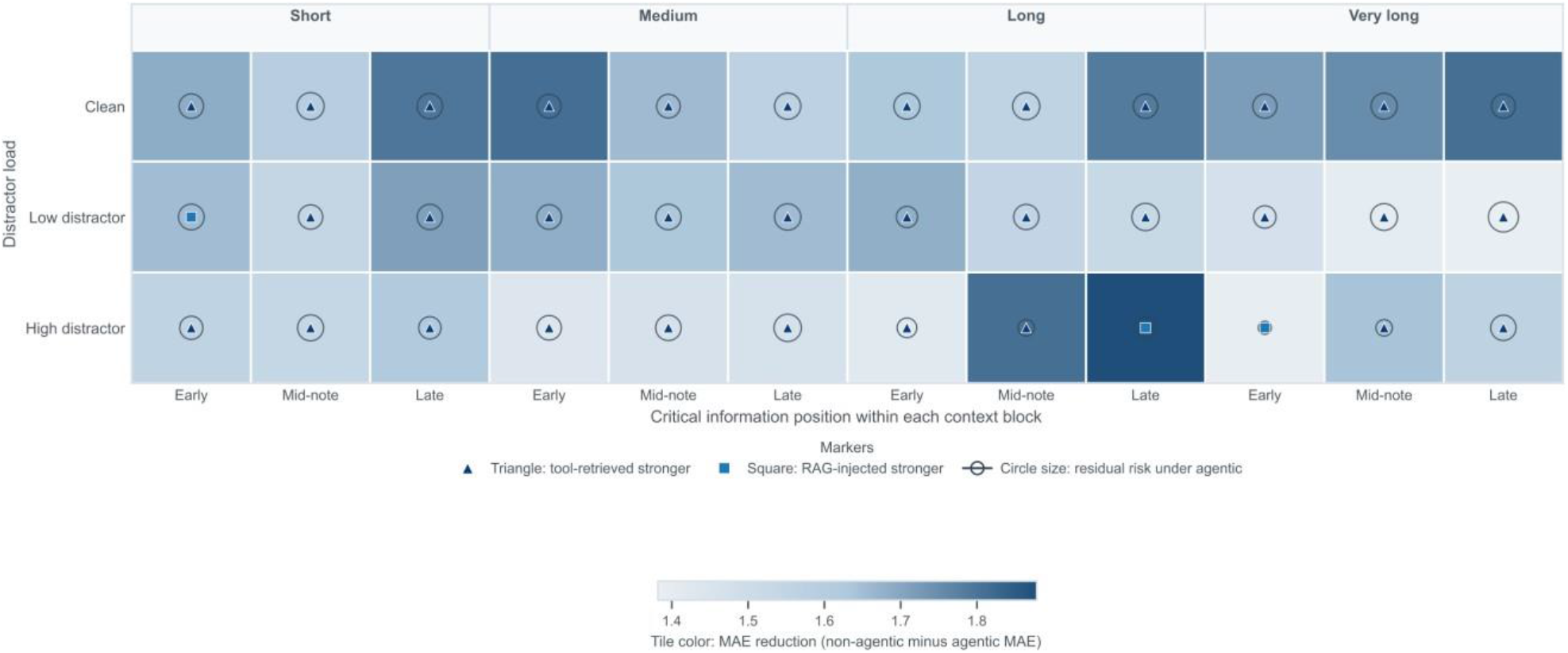
Condition-level RBP gains and residual risk across 36 stress combinations. Heatmap of pooled RBP gain across combinations of context length, distractor load and critical-information position. All combinations showed positive gain (range 1.382–1.878 MAE). Marker shape indicates the dominant framework (triangle: tool-retrieved; square: RAG-injected). Circle size reflects residual risk under agentic workflow. Darker shading indicates larger gain.

### Tool-retrieved pipelines were the dominant protective framework

Framework decomposition showed greater protection for tool-retrieved versus RAG-injected workflows (1.711 versus 1.512 MAE reduction from the non-agentic baseline). Tool-retrieved pipelines were superior in 33 of 36 combinations; RAG-injected pipelines were superior in the remaining 3 (Fig. 4).

**Figure 4.**
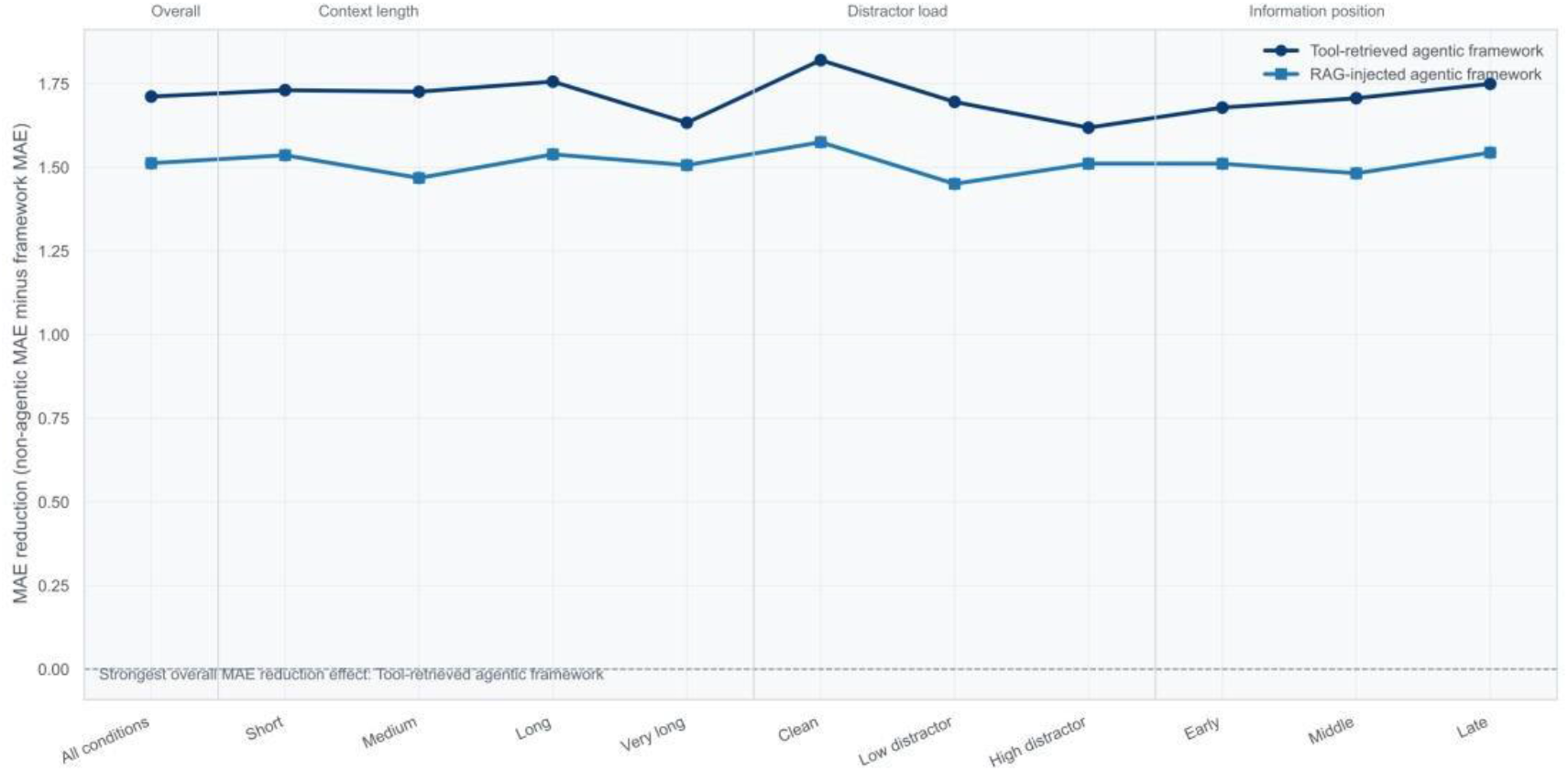
Framework-level decomposition: tool-retrieved versus RAG-injected protection. MAE reduction stratified by context length, distractor load and information position. Tool-retrieved workflows (dark) consistently outperformed RAG-injected workflows (light) across all stress domains.

## Discussion

The central finding of this study is that retrieval workflow design can influence clinical LLM performance as much as model choice. Across all tested combinations of document length, noise and information position, structured retrieval workflows consistently outperformed non-agentic approaches, with an overall reduction in scoring error of 35% (MAE 2.96 versus 4.58; 95% CI for gain 1.57–1.67). These results indicate that structured retrieval improves robustness to chart-context stress, not merely average performance under favourable conditions.

This finding matters because real clinical notes are not clean vignettes. In stroke care, where NIHSS scores directly influence treatment decisions and care-pathway activation, the documentation that clinicians work from is fragmented, lengthy and rarely organised for automated processing. Critical findings may appear late in a note, buried beneath administrative text and repeated entries. When an LLM processes such documentation without structured guidance, degraded performance is not an edge case it is the expected outcome at scale. The relevant safety question for deployed clinical LLMs is therefore not average accuracy on curated benchmarks but reliability under the documentation conditions that define routine care.

This vulnerability extends beyond context length. Prior work has shown that LLMs repeat or elaborate on fabricated clinical details in 50–82% of cases under adversarial conditions, and that prompt-based mitigation reduces but does not eliminate these errors^14^. When presented with authentic versus adversarially modified clinical guidelines, agentic models selected the sham guideline in 40.6% of evaluations, with failure rates exceeding 54% for safety-critical modifications^15^. While our experimental conditions replicate the input structures characteristic of agentic pipelines rather than end-to-end autonomous agents, the results suggest that the retrieval-framing patterns these architectures impose on context are themselves a meaningful source of performance gain. What the present findings add is a complementary dimension: even without adversarial manipulation, realistic documentation structure alone is sufficient to drive systematic performance loss.

The magnitude of benefit was strongly dependent on baseline model capability. Lower-cost models gained far more than frontier models (2.76 versus 0.45 MAE points; differential 2.32; 95% CI 2.22–2.41), suggesting that structured retrieval partially compensates for limited reasoning robustness under context stress. From a deployment perspective, this is particularly relevant because lower-cost models are more likely to be used at scale in resource-constrained settings.

Among structured retrieval approaches, tool-retrieved pipelines outperformed RAG in 33 of 36 combinations. RAG appends retrieved text chunks to the model’s input, reducing the need to scan the full document but still exposing the model to noise and positional artefacts within those chunks^6,11,12^. Tool-retrieved workflows, by contrast, return only the specific information requested, filtering irrelevant content before it enters the reasoning window. The consistent advantage of tool retrieval suggests that reducing unfiltered input, rather than retrieval per se, drives the performance gain.

Structured retrieval did not eliminate risk uniformly. The combination of very long documents, low distractor load and late-positioned critical information retained the highest residual error (MAE 3.22), indicating that positional sensitivity persists when documents are maximally long and target information is deeply embedded. These high-risk stress archetypes are identifiable in advance and represent logical targets for additional quality controls, such as hybrid architectures combining retrieval with clinical rules or knowledge graphs^16^.

Several limitations warrant consideration. The study evaluates a single structured task (NIHSS scoring); whether these findings generalise to less-structured clinical tasks or prospective care settings requires external validation. Model coverage was limited to four Gemini variants, and results may differ across other model families or clinically fine-tuned models. The retrieval implementations represent controlled workflow instantiations rather than production systems, and real deployment introduces additional variability. The primary endpoint was continuous scoring error; whether the observed error reductions translate to fewer misclassifications at actionable clinical thresholds (for example, thrombectomy eligibility) warrants dedicated study.

Beyond stroke scoring, these results suggest that workflow architecture may be a major determinant of LLM reliability under realistic documentation conditions. Lower-cost models, properly scaffolded with structured retrieval, can achieve substantially greater robustness. This matters most where clinical AI is most needed: under-resourced health systems, low- and middle-income settings and high-volume facilities where frontier models are economically or infrastructurally out of reach. Taken together, these findings argue that pre-deployment evaluation of clinical LLMs should stress-test workflow design under realistic chart conditions, and that retrieval architecture may be a more tractable lever for safe, equitable deployment than scaling model capacity alone.

## Methods

### Ethics

This study was approved by the Institutional Review Board of Rambam Medical Center. The requirement for individual informed consent was waived given the retrospective, de-identified nature of the data.

### Study design

We conducted a controlled study to test whether structured retrieval workflows protect LLM performance under context stress during NIHSS scoring. We used 100 de-identified acute stroke cases from the emergency department of a tertiary hospital. Ground-truth NIHSS scores were determined by board-certified physicians based on review of the clinical documentation. For each case, we generated 144 input variants using a fully crossed 4 × 4 × 3 × 3 condition matrix varying four factors: context acquisition method (given, conversational, tool-retrieved, RAG-injected), context length (short, medium, long, very long), distractor load (clean, low noise, high noise) and critical-information position (early, middle, late) (Supplementary Fig. 1).

### Models

We evaluated four Gemini models: Gemini 2.5 Flash-Lite, Gemini 2.5 Flash, Gemini 3 Flash Preview and Gemini 3 Pro Preview. The expected analysis comprised 57,600 model-case-condition runs (4 × 100 × 144). After output parsing and validity filtering, 57,047 runs were retained (99.04%).

### Retrieval workflow definitions

We use the term ‘agentic’ to denote workflows that simulate structured retrieval steps tool-output framing and document injection as implemented in deployed agentic architectures, rather than fully autonomous multi-step reasoning. These conditions mirror the context-formatting patterns that agentic clinical systems produce without requiring the model to autonomously orchestrate retrieval. We implemented four context acquisition modes:

Given (single-pass). A single prompt containing the clinical note followed by NIHSS scoring instructions.

Conversational (history accumulation). The same clinical note presented within a short multi-turn dialogue to emulate information accumulation across a chat history.

Tool-retrieved (tool output). The clinical context returned as a tool-output block (for example, [Tool Results] from <get_patient_info>), followed by NIHSS reference content and the scoring task.

RAG-injected (retrieved documents). The clinical context provided as injected retrieved documents (for example, [Retrieved Documents]), typically including an NIHSS guide and the clinical note, followed by the scoring task.

We grouped these into two workflow classes: non-agentic (given and conversational) and agentic (tool-retrieved and RAG-injected). The primary outcome was retrieval-based protection (RBP) gain, defined as the paired difference in mean absolute error (MAE) between non-agentic and agentic workflows, where MAE is the absolute difference between predicted and ground-truth NIHSS scores. Positive RBP values indicate lower error under agentic workflows.

### Analysis units and model-class stratification

For paired comparisons, the unit of analysis was model x case x stress combination, where each stress combination was defined by context length x distractor load x information position (36 combinations). Within each unit, we aggregated MAE within workflow class across its two acquisition modes, then paired non-agentic MAE against agentic MAE. This produced 14,256 paired analysis units of 14,400 possible.

Model-class stratification was pre-specified based on the hypothesis that newer-generation models (Gemini 3 family) would outperform older-generation models (Gemini 2.5 family), reflecting the generational divide most relevant to deployment decisions. The resulting MAE ranking confirmed this expectation. Weaker models were Gemini 2.5 Flash-Lite and Gemini 2.5 Flash; stronger models were Gemini 3 Pro Preview and Gemini 3 Flash Preview.

### Outcomes

The primary outcome was paired RBP gain overall and by model class. Secondary outcomes were stratified gain across context length, distractor load, information position and composite stress score, condition-level gain and residual risk across the 36 stress combinations, and framework-specific protection (tool-retrieved versus RAG-injected) against the non-agentic baseline.

The composite stress score was defined as the sum of ordinal ranks across length (short = 0, medium = 1, long = 2, very long = 3), distractor load (clean = 0, low = 1, high = 2) and information position (early = 0, middle = 1, late = 2), ranging from 0 to 7.

### Statistical analysis

We report means, absolute differences and 95% confidence intervals (CIs). CIs for paired gain were estimated by nonparametric bootstrap (4,000 resamples; seed 42). We used nonparametric bootstrap on paired analysis units rather than mixed-effects modelling because our primary inference target is the within-unit treatment effect under identical case, model and stress conditions. The paired design inherently controls for case-level and model-level variability by differencing within each unit, eliminating between-unit confounding without requiring distributional assumptions. Bootstrap estimation provides robust confidence intervals without relying on normality of the gain distribution. All analyses were implemented in Python 3.12.

## Data availability

The datasets generated and analysed during the current study are available from the corresponding author upon reasonable request.

## Code availability

Analysis code is available from the corresponding author upon reasonable request.

## Acknowledgements

This work was supported by internal laboratory funding.

## Author contributions

A.G.: Conceptualisation, Methodology, Formal analysis, Data curation, Writing original draft. M.S.: Formal analysis, Data curation. M.O.: Methodology, Formal analysis, Writing review and editing. K.M.: Data curation. A.A.: Conceptualisation, Writing review and editing. Y.B.: Conceptualisation, Writing review and editing. E.K.: Conceptualisation, Methodology, Writing review and editing. S.S.: Conceptualisation, Methodology, Data curation, Writing original draft, Writing review and editing, Supervision.

## Supplementary Information

**Supplementary Figure 1.**
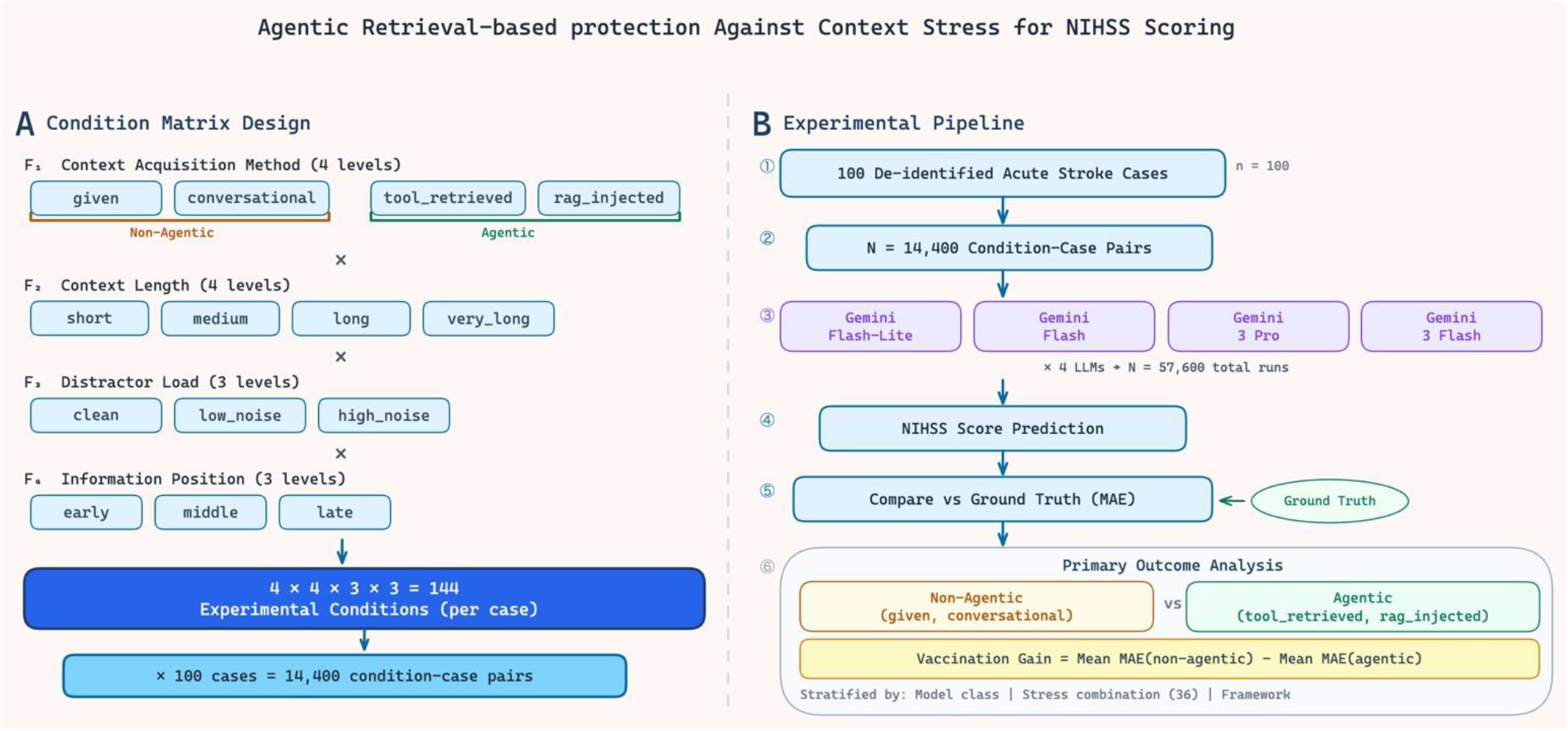
Experimental design. Schematic of the fully crossed 4 × 4 × 3 × 3 condition matrix generating 144 stress conditions for each of 100 de-identified acute stroke cases. The matrix varies context acquisition method (given, conversational, tool-retrieved, RAG-injected), context length (short, medium, long, very long), distractor load (clean, low noise, high noise) and critical-information position (early, middle, late).

